# Incremental Value of Plasma Biomarkers in Predicting Clinical Decline Among Cognitively Unimpaired Older Adults: Results from the A4 trial

**DOI:** 10.1101/2025.07.22.25332015

**Authors:** Babak Khorsand, Elham Ghanbarian, Laura Rabin, Seyed Ahmad Sajjadi, Ali Ezzati

## Abstract

**Objective:** To evaluate the predictive utility of baseline plasma biomarkers and neuropsychological measures in identifying cognitively unimpaired older adults at risk of cognitive and functional decline over five years.

**Background:** The clinical and biological heterogeneity observed in Alzheimer’s disease (AD) complicates design of trials and the identification of appropriate participants. Identifying practical tools to define more homogenous subgroups could enhance clinical trial enrichment and improve early detection.

**Methods:** We analyzed data from the Anti-Amyloid Treatment in Asymptomatic Alzheimer’s Disease (A4) trial and its companion Evaluation of Amyloid Risk and Neurodegeneration (LEARN) observational study. The sample included 866 cognitively unimpaired, amyloid-positive individuals from the A4 trial (comprising participants randomized to receive Solanezumab or placebo) and 343 cognitively unimpaired, amyloid-negative individuals from LEARN. Cognitive/functional decline was defined as an increase of ≥0.5 in Clinical Dementia Rating–Global Score (CDR-GS) during a 240-week period. Using multiple logistic regression models, we evaluated the predictive value of demographic variables, APOE4 status, amyloid PET SUVR, plasma P-tau217, and Alzheimer’s Disease Cooperative Study–Preclinical Alzheimer’s Cognitive Composite (ADCS-PACC) in three groups: A4-Solanezumab, A4-placebo and LEARN. In a sub-study including 656 participants with available data, we assessed the incremental value of additional plasma biomarkers (Aβ42/Aβ40 ratio, GFAP, and NfL).

**Results:** Both plasma P-tau217 and ADCS-PACC significantly improved predictive performance over a base model with demographics and APOE4. The full model combining all predictor variables yielded the highest AUCs across A4 Solanezumab (0.80 ± 0.06), A4 Placebo (0.80 ± 0.06), and LEARN (0.78 ± 0.08). Adding other plasma biomarkers yielded small but consistent improvements in AUC (1–3%).

**Conclusions:** Plasma P-tau217 and ADCS-PACC, individually and in combination, improved prediction of cognitive/functional decline in asymptomatic older adults. Predictive models incorporating these scalable and non-invasive measures improved clinical trial enrichment and earlier identification of at-risk individuals preclinical AD.

## 1. Introduction

Alzheimer’s disease (AD) is the most common neurodegenerative disease, affecting over 55 million people worldwide, with projections indicating a doubling of this figure by 2050 [1]. AD is characterized by a long asymptomatic phase during which neuropathological changes such as amyloid-beta (Aβ) plaque accumulation and tau neurofibrillary tangles may already be present, despite normal cognitive performance [2]. This preclinical stage presents a crucial window for early intervention, particularly with the development of disease-modifying therapies. However, the absence of overt symptoms in this phase poses a major challenge for identifying individuals at highest risk of cognitive decline. This uncertainty complicates both clinical care and trial design, making it essential to identify preclinical individuals most likely to decline over time [3]. Given the heterogeneity in the trajectory of cognitive aging and Alzheimer’s-related decline, it is essential to develop practical, reliable, and scalable tools for risk stratification in asymptomatic populations [4]. This is particularly important for clinical trials targeting early AD, where misclassification of participants can reduce power, inflate costs, and obscure treatment effects.

Several biomarkers have been proposed to aid in the early identification of cognitively unimpaired individuals at elevated risk for Alzheimer’s disease. Amyloid PET imaging, which detects cortical Aβ deposition, has traditionally been used for risk stratification. However, it is costly, not widely available, impractical for large-scale screening, and has shown only modest correlation with longitudinal cognitive decline in asymptomatic individuals. Tau PET imaging demonstrates stronger associations with cognitive performance in older adults and better reflects disease progression, but like amyloid PET, it remains expensive and logistically burdensome[5]. Cerebrospinal fluid (CSF) biomarkers offer diagnostic performance comparable to PET imaging, but their reliance on lumbar puncture makes them less suitable for widespread or repeated clinical use. These limitations have driven increased interest in blood-based biomarkers (BBMs), which are minimally invasive, cost-effective, and scalable for large cohorts. Among these, plasma P-tau217 has shown strong diagnostic and prognostic performance, outperforming other plasma biomarkers in detecting AD pathology and predicting future decline [6-8].

In parallel, cognitive composites such as the Preclinical Alzheimer Cognitive Composite (ADCS-PACC), which integrates sensitive neuropsychological measures across multiple domains, have shown utility in detecting subtle changes in cognition prior to clinical impairment. [9] Together with BBMs, such cognitive composites may offer complementary and scalable tools for risk stratification, particularly in resource-limited clinical care or trial settings.

In this study, we used data from the Anti-Amyloid Treatment in Asymptomatic Alzheimer’s Disease (A4) trial and its companion observational cohort, the Longitudinal Evaluation of Amyloid Risk and Neurodegeneration (LEARN), to examine the predictive utility of baseline features—including plasma P-tau217, ADCS-PACC, APOE4 status, amyloid PET SUVR, and demographics—in predicting cognitive/functional decline over five years. We employed a logistic regression modeling approach to compare individual and joint contributions of each predictor across three well-characterized study arms: A4 solanezumab-treated, A4 placebo, and LEARN. In a secondary analysis we assessed the added value of plasma Aβ42/Aβ40, GFAP, and NfL in enhancing predictive performance in the subset of participants with available plasma data. Importantly, our goal was to evaluate the combined predictive utility of these measures, considering their performance in integrated models rather than in isolation, across both amyloid-positive and amyloid-negative individuals. By identifying a parsimonious and scalable feature set, we aim to inform early detection strategies and enhance clinical trial enrichment.

## 2. Methods

### 2.1 Design and Participants

We used data from the Anti-Amyloid Treatment in Asymptomatic Alzheimer’s Disease (A4) and Longitudinal Evaluation of Amyloid Risk and Neurodegeneration (LEARN) studies. The A4 study was a 240 week, double-blind, and placebo-controlled randomized clinical trial designed to evaluate the efficacy of solanezumab in slowing cognitive decline. A4 enrolled 1169 healthy adults aged 65 to 85, all with Clinical Dementia Rating (CDR) score of 0, Mini-Mental State Examination (MMSE) of 25-30, and Logical Memory delayed Recall (LMDR IIA) score of 6-18, who were amyloid-positive via PET imaging as described below [10, 11]. The LEARN study, an observational arm of A4 study, enrolled 538 cognitively unimpaired individuals who met the same screening criteria as A4 participants except for amyloid status, and were classified as amyloid-negative on PET imaging.

This analysis included participants from three study arms: the A4 Solanezumab (N=578), A4 Placebo (N=591), and LEARN (N=538) who had CDR scores at baseline and at least two follow-up visits. For the primary analysis, we included participants with available data on demographics, APOE4 genotype, ADCS-PACC, plasma P-tau217, and final CDR score: A4 Solanezumab (N=429), A4 Placebo (N=437), and LEARN (N=343). In a secondary analysis, we examined the added value of other blood-based biomarkers (BBMs). This sub-study included participants with plasma data for Aβ42, Aβ40, GFAP, and NfL at screening: A4 Solanezumab (N=187), A4 Placebo (N=179), an LEARN (N=290). Figure 1 provides an overview of participant flow and sample sizes across analyses.

**Figure 1.**
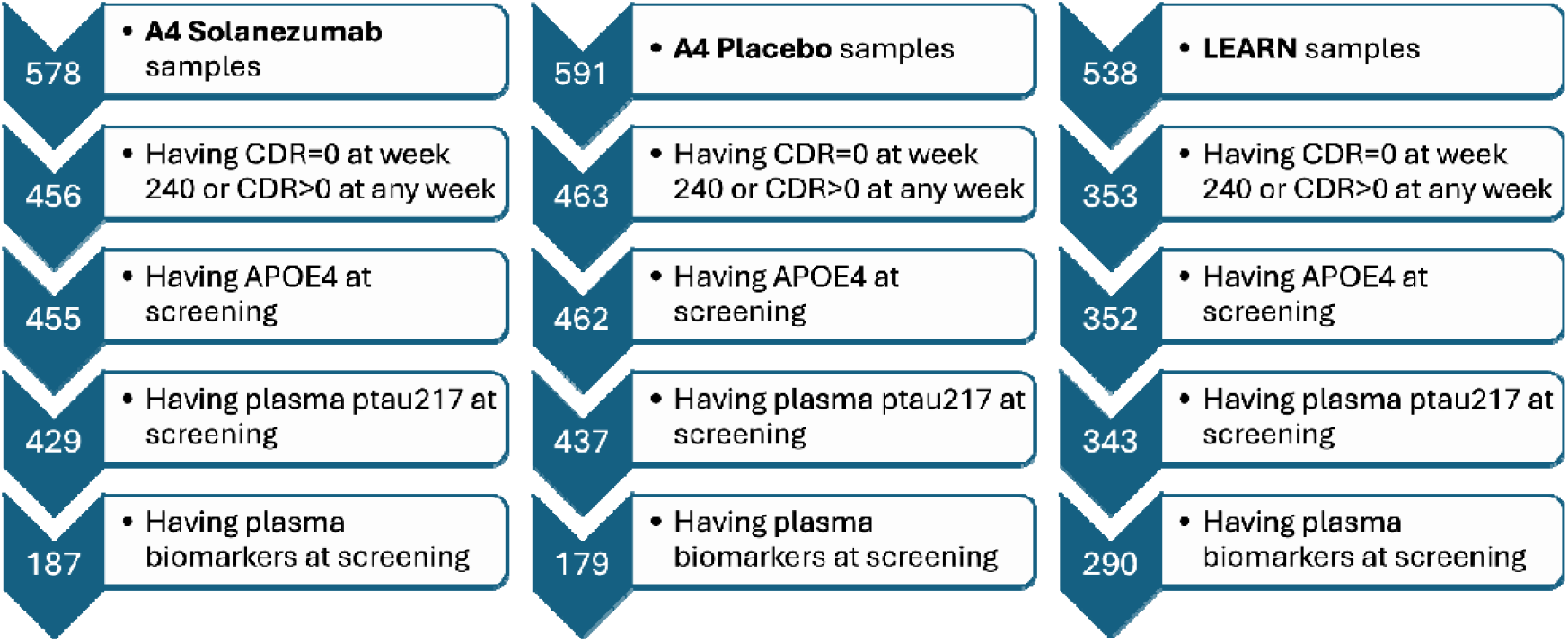
Flow chart of study participants.

### 2.2 Study Measures

#### Clinical outcomes

The primary cognitive/function outcome was the Clinical Dementia Rating-Global Score (CDR-GS) scale which involves semi structured interviews with both the participant and an informant to gather information required to evaluate cognitive functioning across six key areas: memory, orientation, judgment and problem-solving, involvement in community activities, home and leisure pursuits, and personal care. Each area is rated independently, using a five-point scale to reflect the severity of impairment: 0 (no impairment), 0.5 (very mild/questionable), (mild), 2 (moderate), and 3 (severe dementia) [12]. Our outcome variable was cognitive/functional decline defined as an increase of ≥0.5 in CDR-GS over the course of the 240 weeks.

#### Cognitive measures

The Preclinical Alzheimer Cognitive Composite (ADCS-PACC) is a composite score derived from standardized neuropsychological tests that assess memory, executive function, and global cognition. The ADCS-PACC score was calculated by summing z-scores from the following assessments

- Mini-Mental State Examination (MMSE): A screening tool evaluating global cognition, scored on a scale from 0 to 30
- Delayed Logical Memory (DLM): Measures the ability to recall a narrative 15 minutes after the initial recall, with a score range of 0 to 25
- Digit Symbol Substitution (DSS): Assesses executive function, processing speed, and working memory, with a maximum possible score of 91
- Free and Cued Selective Reminding Test (FCSRT): A multi-trial word recall assessment, where scores include free recall (FR) alone (ranging from 0 to 48) and total recall (TR96), which sums FR and cued recall (ranging from 0 to 96).

#### Imaging Biomarkers

Amyloid burden was quantified using 18F-florbetapir PET imaging. Quantitative measures were derived using the standardized uptake value ratio (SUVR), calculated by comparing tracer uptake in target cortical regions to a reference region. A threshold of SUVR>1.15, or SUVR between 1.1-1.15 with concordant visual read, was used to classify as amyloid-positive. Those below this threshold without visual read support were considered as amyloid-negative [11].

#### Plasma biomarkers

Plasma P-tau217, reflecting tau protein phosphorylated at threonine 217, was measured from blood samples using an analytically validated electrochemiluminescence (ECL) immunoassay platform [7]. Additional plasma biomarkers analyzed in a sub-study included amyloid beta 42/40 ratio (Aβ42/Aβ40), glial fibrillary acidic protein (GFAP), and neurofilament light chain (NfL), all quantified using ultrasensitive single molecule array (Simoa) assays [13]. All measurements were obtained from plasma samples collected under standardized conditions at baseline.

#### APOE4 Genotyping

APOE ε4 carrier status was determined using genomic DNA extracted from blood samples collected at baseline. Genotyping was performed using standardized SNP-based assays to detect the presence of the ε4 allele. Participants were classified as APOE4 carriers if they had at least one ε4 allele (heterozygous or homozygous), and as non-carriers otherwise.

### 2.3 Statistical analysis

Participants were stratified into three study arms: A4 Solanezumab, A4 Placebo (both amyloid-positive), and LEARN (amyloid-negative). Within each group, participants were categorized as *Decliners* or *Stables* based on longitudinal changes in CDR-GS. Decliners were defined as individuals whose CDR global score increased by ≥0.5 points at two or more consecutive follow-up visits; Stables were those who maintained a CDR global score of 0 at the final visit (week 240).

Descriptive statistics were used to compare baseline characteristics between Decliners and Stables within each study arm. Normality of continuous variables was assessed using the Shapiro–Wilk test. Group differences in continuous variables with a normal distribution were tested using independent sample t-tests, while non-normally distributed continuous variables were tested using Mann-Whitney U test. Categorical variables were compared using chi-square tests.

To predict cognitive outcome status (Decliner vs. Stable), a series of logistic regression models were developed using different combinations of baseline predictors. The primary eight models included:

- **Base Model:** Demographic variables (age, sex, education) and APOE4 carrier status
- **+Amyloid Model:** Base Model plus amyloid PET SUVR
- **+P-tau217 Model:** Base Model plus plasma P-tau217
- **+ADCS-PACC Model:** Base Model plus ADCS-PACC score
- **+Amyloid & P-tau217 Model:** Base Model plus both amyloid PET SUVR and P-tau217
- **+Amyloid & ADCS-PACC Model:** Base Model plus both amyloid PET SUVR and ADCS-PACC
- **+P-tau217 & ADCS-PACC Model:** Base Model plus both P-tau217 and ADCS-PACC
- **Full Model:** Base Model plus all three additional predictors (amyloid PET SUVR, P-tau217, and ADCS-PACC)

Each model was trained separately within the Solanezumab, Placebo, and LEARN arms. Model performance was assessed using five-fold cross-validation, and mean area under the ROC curve (AUC), sensitivity, specificity, positive predictive value (PPV), and negative predictive value (NPV) were calculated. Standard deviations across folds were used to summarize variability in AUC estimates.

In a secondary analysis, the effect of adding other BBMs—Aβ42/Aβ40 ratio, GFAP, and NfL—was evaluated in a subset of participants with available plasma data. BBM variables were added to each of the eight primary models to form eight augmented models. The impact of BBM inclusion on model fit was tested using Likelihood Ratio Tests, and changes in cross-validated AUC were reported.

All analyses were performed in R version 4.4.0, with two-sided p-value < 0.05 considered statistically significant.

## 3. Results

### 3.1 Sample characteristics

The overall sample had a mean age of 71.4 ± 4.6 years; 58.7% were female, and 49.1% were APOE4 carriers. Based on longitudinal CDR-GS data, 183 participants (42.7%) in the A4 Solanezumab group, 170 (38.9%) in A4 Placebo, and 90 (26.2%) in LEARN were classified as Decliners. The remaining participants were categorized as Stables.

Across all three arms, Decliners had significantly higher baseline P-tau217 levels and lower ADCS-PACC scores (p < 0.01). Differences in other variables, including age and APOE4 frequency, varied by arm and are summarized in Table 1.

**Table 1.**
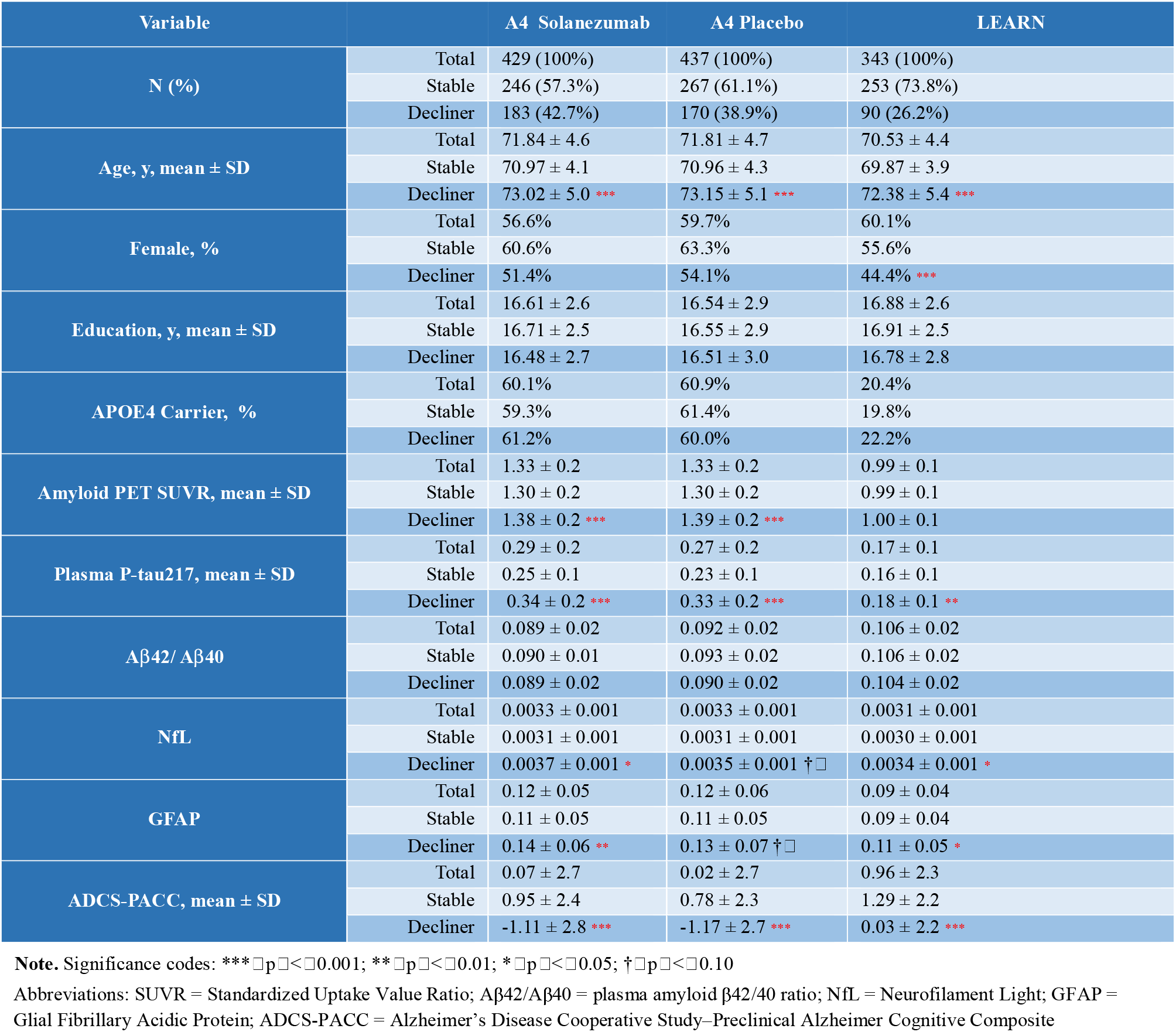
Baseline Characteristics of Participants Across Study Arms.

### 3.2 Prediction of the cognitive outcome

We developed and validated eight logistic regression models using different combinations of baseline predictors and evaluated their performance using five-fold cross-validation (Table 2).

**Table 2.**
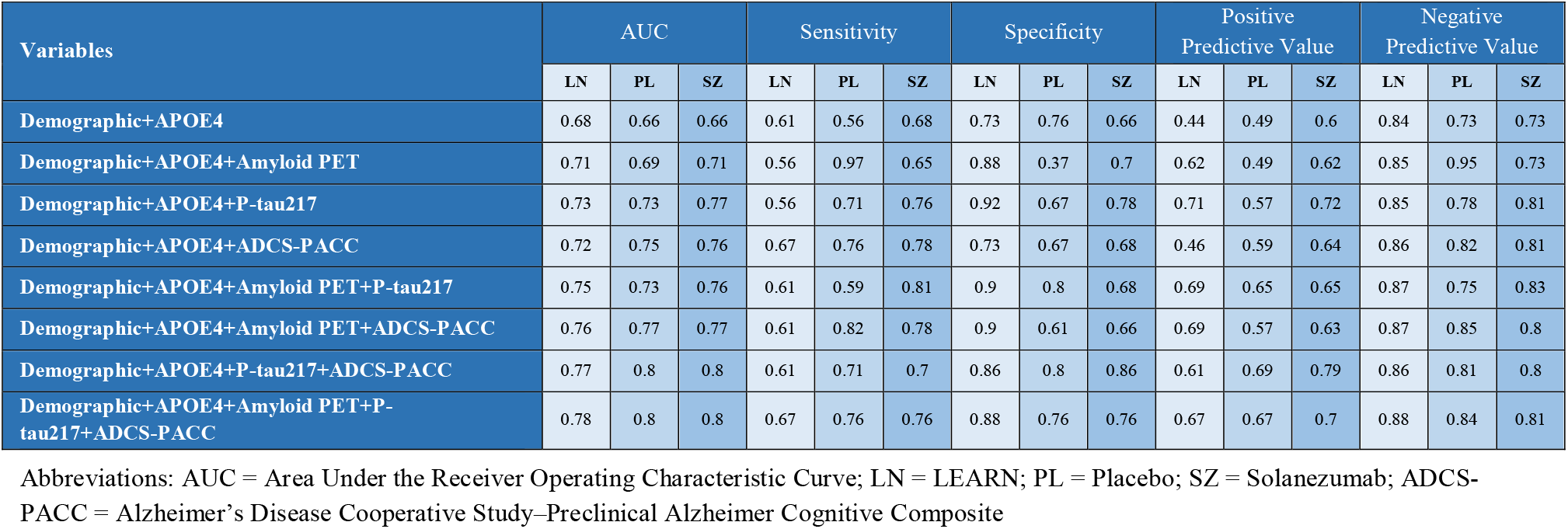
Diagnostic Performance Metrics for Predictive Models Across Study Arms.

| Variables | AUC |  |  | Sensitivity |  |  | Specificity |  |  | Positive Predictive Value |  |  | Negative Predictive Value |  |  |
| --- | --- | --- | --- | --- | --- | --- | --- | --- | --- | --- | --- | --- | --- | --- | --- |
|  | LN | PL | SZ | LN | PL | SZ | LN | PL | SZ | LN | PL | SZ | LN | PL | SZ |
| Demographic+APOE4 | 0.68 | 0.66 | 0.66 | 0.61 | 0.56 | 0.68 | 0.73 | 0.76 | 0.66 | 0.44 | 0.49 | 0.6 | 0.84 | 0.73 | 0.73 |
| Demographic+APOE4+Amyloid PET | 0.71 | 0.69 | 0.71 | 0.56 | 0.97 | 0.65 | 0.88 | 0.37 | 0.7 | 0.62 | 0.49 | 0.62 | 0.85 | 0.95 | 0.73 |
| Demographic+APOE4+P-tau217 | 0.73 | 0.73 | 0.77 | 0.56 | 0.71 | 0.76 | 0.92 | 0.67 | 0.78 | 0.71 | 0.57 | 0.72 | 0.85 | 0.78 | 0.81 |
| Demographic+APOE4+ADCS-PACC | 0.72 | 0.75 | 0.76 | 0.67 | 0.76 | 0.78 | 0.73 | 0.67 | 0.68 | 0.46 | 0.59 | 0.64 | 0.86 | 0.82 | 0.81 |
| Demographic+APOE4+Amyloid PET+P-tau217 | 0.75 | 0.73 | 0.76 | 0.61 | 0.59 | 0.81 | 0.9 | 0.8 | 0.68 | 0.69 | 0.65 | 0.65 | 0.87 | 0.75 | 0.83 |
| Demographic+APOE4+Amyloid PET+ADCS-PACC | 0.76 | 0.77 | 0.77 | 0.61 | 0.82 | 0.78 | 0.9 | 0.61 | 0.66 | 0.69 | 0.57 | 0.63 | 0.87 | 0.85 | 0.8 |
| Demographic+APOE4+P-tau217+ADCS-PACC | 0.77 | 0.8 | 0.8 | 0.61 | 0.71 | 0.7 | 0.86 | 0.8 | 0.86 | 0.61 | 0.69 | 0.79 | 0.86 | 0.81 | 0.8 |
| Demographic+APOE4+Amyloid PET+P-tau217+ADCS-PACC | 0.78 | 0.8 | 0.8 | 0.67 | 0.76 | 0.76 | 0.88 | 0.76 | 0.76 | 0.67 | 0.67 | 0.7 | 0.88 | 0.84 | 0.81 |

Abbreviations: AUC = Area Under the Receiver Operating Characteristic Curve; LN = LEARN; PL = Placebo; SZ = Solanezumab; ADCS-PACC = Alzheimer's Disease Cooperative Study–Preclinical Alzheimer Cognitive Composite

In the Placebo arm of A4 Study, the base model, which included demographics and APOE4 status, achieved an overall AUC of 0.66 ± 0.06. Adding amyloid PET SUVR increased the AUC to 0.69 ± 0.06, but this improvement was not statistically significant (ΔAUC = 0.03, p = 0.13). By contrast, the addition of plasma P-tau217 or ADCS-PACC significantly enhanced model performance. The P-tau217 model achieved an AUC of 0.73 ± 0.06 (ΔAUC = 0.07 vs. base, p < 0.001), and the ADCS-PACC model achieved an AUC of 0.75 ± 0.06 (ΔAUC = 0.09 vs. base, p < 0.001). Both models significantly outperformed the amyloid PET model (P-tau217 vs. amyloid PET: p < 0.05; ADCS-PACC vs. amyloid PET: p < 0.01). The model combining both P-tau217 and ADCS-PACC achieved the highest predictive performance among dual-feature models, with an AUC of 0.80 ± 0.06 (ΔAUC = 0.14 vs. base, p < 0.001). Including amyloid PET SUVR in the full model did not improve performance, as the AUC remained at 0.80 ± 0.06. Detailed diagnostic performance metrics—including sensitivity, specificity, PPV, and NPV—for each model across the three study arms are reported in Table 2. These patterns were replicated in the A4 Solanezumab and LEARN arms, with cross-validated AUC values for all eight models presented in Figure 2. The corresponding pairwise model comparisons is summarized in Table 3.

**Table 3.** Pairwise Differences in Model Performance (AUC) Across Study Arms with Incremental Addition of Predictors.

| vs. Model ↓ / Model → |  | Base | +Amyloid | +P-tau217 | +ADCS-PACC | P-tau217+ADCS-PACC |
| --- | --- | --- | --- | --- | --- | --- |
| **LEARN** | Base | — | 0.03 (>0.05) | **0.05 (<0.05)** | **0.04 (<0.05)** | **0.09 (<0.001)** |
|  | +Amyloid |  | — | **0.04 (<0.05)** | **0.05 (<0.05)** | **0.07 (<0.001)** |
|  | +P-tau217 |  | 0.02 (>0.05) | — | **0.04 (<0.05)** | **0.04 (<0.05)** |
|  | +ADCS-PACC |  | **0.04 (<0.05)** | **0.05 (<0.05)** | — | **0.05 (<0.05)** |
| **A4 Placebo** | Base | — | 0.03 (>0.05) | **0.07 (<0.001)** | **0.09 (<0.001)** | **0.14 (<0.001)** |
|  | +Amyloid |  | — | **0.04 (<0.05)** | **0.06 (<0.05)** | **0.11 (<0.001)** |
|  | +P-tau217 |  | 0 | — | **0.07 (<0.001)** | **0.07 (<0.001)** |
|  | +ADCS-PACC |  | 0.02 (>0.05) | 0.03 (>0.05) | — | 0.03 (>0.05) |
| **A4 Solanezumab** | Base | — | 0.05 (<0.05) | **0.11 (<0.001)** | **0.10 (<0.001)** | **0.14 (<0.001)** |
|  | +Amyloid |  | — | **0.06 (<0.05)** | **0.05 (<0.05)** | **0.09 (<0.001)** |
|  | +P-tau217 |  | 0 | — | 0.03 (>0.05) | 0.03 (>0.05) |
|  | +ADCS-PACC |  | 0.01 (>0.05) | **0.04 (<0.05)** | — | **0.04 (<0.05)** |

**Note.** Differences in cross-validated area under the ROC curve (AUC) between logistic regression models including various combinations of baseline predictors: demographics + APOE4 (Base), amyloid PET SUVR (+Amyloid), plasma P-tau217, ADCS-PACC , and their combinations. Values represent ΔAUC with associated significance levels. Abbreviations: AUC = Area Under the Curve; SUVR = Standardized Uptake Value Ratio; ADCS-PACC = Alzheimer’s Disease Cooperative Study–Preclinical Alzheimer Cognitive Composite.

**Figure 2.**
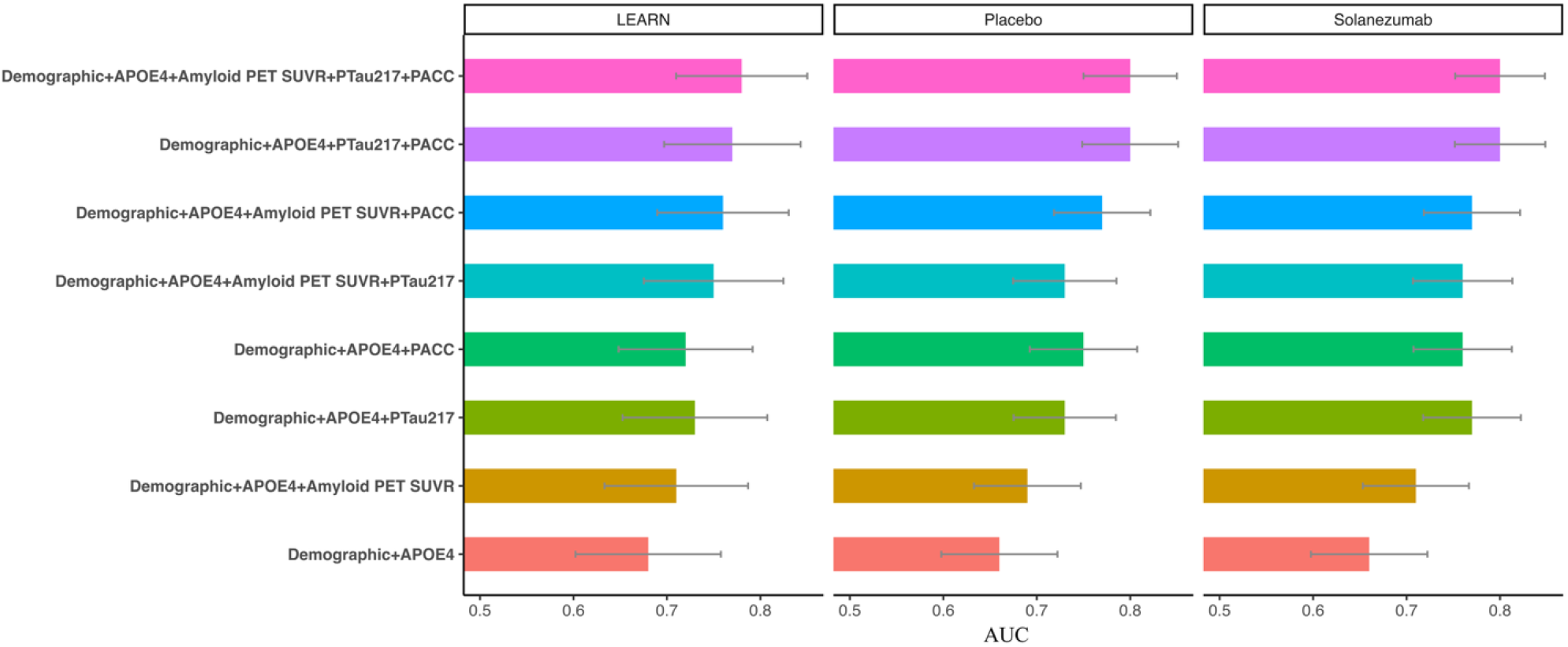
Comparative Model Performance (AUC) for Predicting Cognitive Decline Across Study Arms Bar plots showing mean cross-validated AUC values (with standard error bars) for logistic regression models predicting cognitive/functional Decliners in the LEARN, A4 Placebo, and A4 Solanezumab groups. Models incorporate combinations of demographics, APOE4, amyloid PET SUVR, plasma P-tau217, and PACC. Abbreviations: AUC = Area Under the Receiver Operating Characteristic Curve; PACC = Alzheimer’s Disease Cooperative Study–Preclinical Alzheimer Cognitive Composite; SUVR = Standardized Uptake Value Ratio.

**Figure 3.**
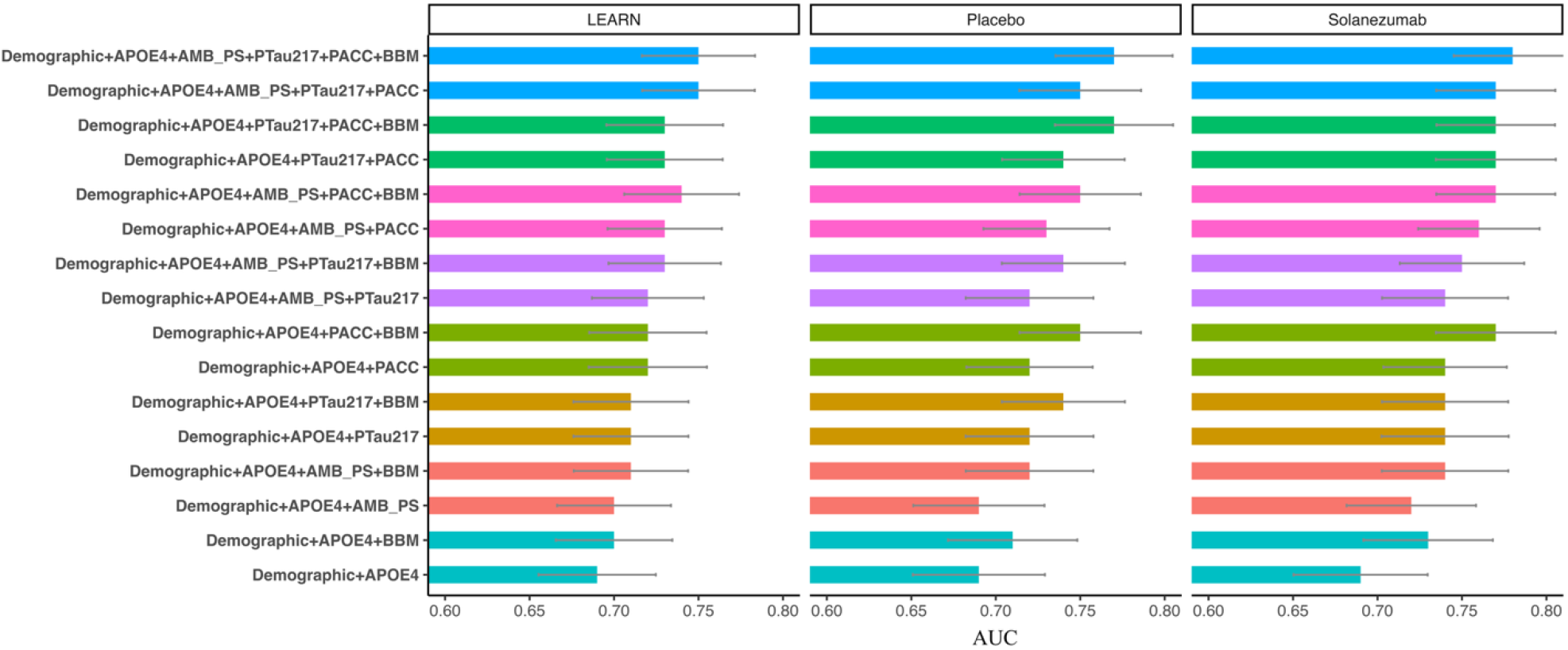
Logistic regression model performance for each set of independent variables in predicting Decliners in the presence of blood-based markers (BBM) including plasma Aβ42/Aβ40 ratio, plasma GFAP, and plasma NfL in addition to variables considered in the main study. Abbreviations: PACC = Alzheimer’s Disease Cooperative Study–Preclinical Alzheimer Cognitive Composite; AMB_PS = Amyloid Beta Positron Emission Tomography Standardized Uptake Value Ratio.

### 3.3. Sub-Study: Incremental Predictive Value of other BBMs

To assess the added predictive value of other blood-based biomarkers (BBMs), we conducted a secondary analysis in a subset of 656 participants with available plasma data for Aβ42/Aβ40 ratio, GFAP, and NfL. Each of the eight original models was re-estimated with BBMs included as additional predictors.

Across all study arms, BBMs inclusion modestly improved AUC values. The greatest relative improvement (~4%) was observed when BBMs were added to the base model, increasing the AUC from 0.69 to 0.73 in A4 Solanezumab arm. Similar gains (~2–3%) were seen when BBMs were added to single-feature models that included only amyloid PET, P-tau217, or ADCS-PACC. For models that already included two predictors, BBMs improved AUC by approximately 1–2%. For the full model, which already incorporated demographics, APOE4, amyloid PET SUVR, P-tau217, and ADCS-PACC, addition of BBMs showed the smallest improvement, with AUC increasing by only ~1%.

In most cases, these AUC gains were small and not statistically significant (DeLong p > 0.05, Supplementary Table 1), suggesting diminishing returns from adding BBMs to models that already include plasma P-tau217 and cognitive measures.

## 4. Discussion

This study provides a comparative evaluation of predictive models incorporating demographics, plasma biomarkers, a cognitive composite score, and imaging features to forecast cognitive/functional decline, as measured by the CDR-GS in cognitively unimpaired older adults. Our findings demonstrate that models including plasma P-tau217 and the ADCS-PACC outperform those using either biomarker alone, and showed stronger predictive accuracy than models relying on amyloid PET SUVR. In particular, the combination of P-tau217 and ADCS-PACC yielded the best discrimination between Decliners and Stables across all three study arms, including amyloid-positive and amyloid-negative individuals. The limited incremental value of adding amyloid PET SUVR once P-tau217 and ADCS-PACC were included highlights the practicality of relying on these scalable and minimally invasive measures for early risk stratification.

Our results are consistent with prior studies establishing P-tau217 as a sensitive and specific plasma biomarker for AD pathology and progression. Palmqvist et al. demonstrated that plasma P-tau217 outperformed other plasma markers, such as P-tau181 and NfL, in distinguishing clinically and neuropathologically defined AD from other neurodegenerative conditions [14]. Also, they demonstrated a higher diagnostic accuracy of plasma P-tau217 for clinical AD in comparison with other plasma biomarkers such as P-Tau181 and NfL [14]. Similarly, Karikari et al. showed that plasma P-tau levels effectively differentiated AD from cognitively normal controls [15] and other studies have also reported enhanced prognostic accuracy when combining plasma P-tau and NfL with demographics and APOE ε4 status in individuals with mild cognitive impairment [16]. Our study extends these findings to the preclinical stage, showing that P-tau217 remains a strong predictor of decline even among individuals without cognitive impairment at baseline.

The ADCS-PACC, originally developed to capture subtle cognitive change in preclinical AD, also demonstrated strong prognostic value. Our findings reaffirm its sensitivity in detecting early decline, even among individuals who are clinically normal at baseline [17]. Notably, both P-tau217 and ADCS-PACC differentiated Decliners from Stables in the LEARN cohort, which included only amyloid-negative individuals. This suggests these markers might be sensitive to early neurodegenerative processes or cognitive changes that are not solely dependent on cortical amyloidosis reaching conventional positivity thresholds, or perhaps reflect very early stages of amyloid pathology not yet meeting the full criteria for positivity[18-20]. This warrants further investigation into the underlying biological pathways in these amyloid-negative individuals who still experience decline, potentially involving co-pathologies or alternative mechanisms.

Previous research has explored the utility of plasma biomarkers and cognitive composites in forecasting cognitive decline among older adults. Papp et al. examined the predictive value of plasma P-tau217 and ADCS-PACC in the A4 cohort and found that these measures were associated with longitudinal cognitive outcomes, though their analyses were limited to the active treatment arm and did not compare predictive models across study groups or evaluate the incremental benefit of combining biomarkers with cognitive composites in integrated models [21]. Similarly, Bransby et al. demonstrated that baseline ADCS-PACC scores could predict subsequent cognitive trajectories in preclinical AD, but they did not incorporate plasma biomarkers into their predictive models [22]. Our study builds on and extends these findings by systematically comparing the predictive utility of plasma P-tau217, ADCS-PACC, and other blood-based markers—both individually and in combination—across three well-characterized cohorts (A4 Solanezumab, A4 Placebo, and LEARN). By evaluating the additive and comparative value of scalable, non-invasive markers in diverse participant groups, we identify practical strategies for risk stratification in both research and clinical settings.

In our sub-study evaluating additional blood-based biomarkers (Aβ42/Aβ40, GFAP, and NfL), we observed modest improvements in model performance when these markers were added to simpler models. However, once P-tau217 and ADCS-PACC were included, the incremental value of GFAP and NfL was minimal, suggesting potential redundancy or overlapping information. This finding is consistent with prior literature indicating that while GFAP and NfL reflect neuroinflammation and neuroaxonal injury, respectively, their predictive value may be most apparent in symptomatic or mixed pathology cohorts[23, 24]. It is possible that these markers may help define biological subtypes or endophenotypes in future work, even if their additive value to simple predictive models is limited [25].

The implications of these findings are significant for both AD research and clinical practice. The ability to identify at-risk cognitively unimpaired individuals using scalable, non-invasive, and cost-effective measures like plasma P-tau217 and ADCS-PACC can support more efficient enrichment of preclinical AD trials, improving power, reducing costs, and minimizing exposure of low-risk individuals to unnecessary interventions. As disease-modifying therapies become available, predictive tools may also help identify individuals most likely to benefit from early intervention. Our findings support a pragmatic model of risk prediction using a small number of high-yield measures, consistent with the growing emphasis on parsimony and accessibility in AD research and care.

Our findings also support the principle of parsimony in predictive modeling. Although the full model achieved the highest AUC, simpler models that included only demographics, APOE4, plasma P-tau217, and ADCS-PACC performed nearly as well. This suggests that a small, well-selected feature set may offer high predictive utility while minimizing participant burden and analytical complexity. Such streamlined models are well-suited for practical implementation in both research and clinical settings [17].

Several study limitations should be noted. First, the sub-study evaluating additional BBMs had a smaller sample size, limiting power to detect small effects and potentially biasing comparisons. Second, while the ADCS-PACC is a well-validated composite, the inclusion of additional cognitive tasks (e.g., episodic memory binding or digital tasks) might further enhance sensitivity and specificity. Third, we relied on baseline measures only; incorporating longitudinal trajectories of plasma and cognitive markers may improve predictive performance of the models further. Finally, our findings should be validated in more diverse cohorts, including those with broader racial/ethnic representation and greater variability in comorbidities.

In conclusion, this study provides strong support for the use of plasma P-tau217 and ADCS-PACC as independent and complementary predictors of cognitive and functional decline in cognitively unimpaired individuals. Models incorporating these readily accessible and non-invasive measures offer a promising avenue for improving participant selection in prevention trials and facilitating earlier intervention strategies in preclinical AD. Future research should evaluate the generalizability of these models in more diverse populations, assess the added value of longitudinal markers, and further investigate biological pathways in individuals who decline despite amyloid-negative status at baseline.

## Data Availability

All data produced in the present study are available upon reasonable request to the authors

https://www.a4studydata.org/

## Author Contributions

Babak Khorsand (Conceptualization; Methodology; Writing - Original Draft; Formal analysis; Investigation); Elham Ghanbarian (Writing - Review & Editing); Laura Rabin (Writing - Review & Editing; Conceptualization); Seyed Ahmad Sajjadi (Writing - Review & Editing; Formal analysis); Ali Ezzati (Writing - Review & Editing; Formal analysis; Investigation; Conceptualization).

## Acknowledgements

This study was supported in part by grants from the National Institute of Health (NIA K23 AG063993; R01 AG080635); the Alzheimer’s Association (SG-24-988292 ISAVRAD); Cure Alzheimer’s Fund.

## Source of data and Funding

The A4 Study is a secondary prevention trial in preclinical Alzheimer’s disease, aiming to slow cognitive decline associated with brain amyloid accumulation in clinically normal older individuals. The A4 Study is funded by a public-private philanthropic partnership, including funding from the National Institutes of Health-National Institute on Aging (U19 AG010483, U24AG057437, R01 AG063689), Eli Lilly and Company, Alzheimer’s Association, Accelerating Medicines Partnership, GHR Foundation, an anonymous foundation and additional private donors, with in-kind support from Avid and Cogstate. The companion observational Longitudinal Evaluation of Amyloid Risk and Neurodegeneration (LEARN) Study is funded by the Alzheimer’s Association (LEARN-15-338729) and GHR Foundation. The A4 and LEARN Studies are led by Dr. Reisa Sperling at Brigham and Women’s Hospital, Harvard Medical School and Dr. Paul Aisen at the Alzheimer’s Therapeutic Research Institute (ATRI), University of Southern California. The A4 and LEARN Studies are coordinated by ATRI at the University of Southern California, and the data are made available through the Laboratory for Neuro Imaging at the University of Southern California. The participants screening for the A4 Study provided permission to share their de-identified data to advance the quest to find a successful treatment for Alzheimer’s disease.

---

**Supplementary Table 1.** Incremental improvement in model performance (AUC) for adding BBM to models across study arms (LEARN, Placebo, and Solanezumab).

| Models | +BBM |  |  |
| --- | --- | --- | --- |
|  | LEARN | A4 Placebo | A4 Solanezumab |
| Base | 0.01 (>0.05) | 0.02 (>0.05) | 0.04 (>0.05) |
| Base + Amyloid | 0.01 (>0.05) | 0.03 (>0.05) | 0.02 (>0.05) |
| Base + P-tau217 | <0.01 (>0.05) | 0.02 (>0.05) | <0.01 (>0.05) |
| Base + ADCS-PACC | <0.01 (>0.05) | 0.03 (>0.05) | 0.03 (>0.05) |
| Base + Amyloid + P-tau217 | 0.01 (>0.05) | 0.02 (>0.05) | 0.01 (>0.05) |
| Base + Amyloid + ADCS-PACC | 0.01 (>0.05) | 0.02 (>0.05) | 0.01 (>0.05) |
| Base + P-tau217 + ADCS-PACC | <0.01 (>0.05) | 0.03 (>0.05) | <0.01 (>0.05) |
| Base + Amyloid + P-tau217 + ADCS-PACC | <0.01 (>0.05) | 0.02 (>0.05) | 0.01 (>0.05) |

Note. Abbreviations: AUC = Area Under the Receiver Operating Characteristic Curve; BBM = Blood-Based Biomarker; ADCS-PACC = Alzheimer's Disease Cooperative Study–Preclinical Alzheimer Cognitive Composite.

## Notes

### Competing Interest Statement

The authors have declared no competing interest.

### Author Declarations

https://www.a4studydata.org/ The datasets used in our study were aggregated/summary data, and all individual-level data had been de-identified prior to its use in this study.

